# COVID-19 transmission in the U.S. before vs. after relaxation of state social distancing measures

**DOI:** 10.1101/2020.07.15.20154534

**Authors:** Alexander C. Tsai, Guy Harling, Zahra Reynolds, Rebecca F. Gilbert, Mark J. Siedner

## Abstract

**Background:** Weeks after issuing social distancing orders, all U.S. states and the District of Columbia at least partially relaxed these measures. Critical unanswered questions remain about the timing of relaxation, and if and how unregulated social distancing measures can be sustained while effectively maintaining epidemic control.

**Methods:** We identified all statewide social distancing measures that were implemented and/or relaxed in the U.S. between March 10-July 15, 2020, triangulating data from state government and third-party sources. Using segmented linear regression, we evaluated the extent to which social distancing measure relaxation affected epidemic control, as indicated by the time-varying, state-specific effective reproduction number (R*_t_*).

**Results:** In the eight weeks prior to relaxation, mean R*_t_* declined by 0.012 units per day (95% CI, -0.013 to -0.012), and 46/51 jurisdictions achieved R*_t_* < 1.0 by the date of relaxation. After relaxation of social distancing, R*_t_* reversed course and began increasing by 0.007 units per day (95% CI, 0.006-0.007), reaching a mean R*_t_* of 1.16 eight weeks later, with only 9/51 jurisdictions maintaining R*_t_* <1.0. Indicators often used to motivate relaxation at the time of relaxation (e.g. test positivity rate <5%) predicted greater post-relaxation epidemic growth.

**Conclusions:** We detected an immediate and significant reversal in epidemic growth gains after relaxation of social distancing measures across the U.S. These results illustrate the potential pitfalls of premature relaxation of social distancing measures in the U.S.

## Introduction

The U.S. is home to the largest epidemic of severe acute respiratory syndrome coronavirus 2 (SARS-CoV-2) globally, having surpassed 3 million reported cases and 140,000 deaths by early July [1]. After experiencing large, localized epidemics in March and April, all fifty U.S. states and the District of Columbia implemented social distancing measures in an attempt to interrupt transmission and reduce morbidity and mortality from coronavirus disease 2019 (COVID-19). Several studies from the U.S. [2-5] and elsewhere [2, 6, 7] have demonstrated the effectiveness of social distancing measures in reducing COVID-19 case growth and the resulting morbidity and mortality.

Concerns about adverse economic, population health, and social spillover consequences of social distancing [8-11] have undermined adherence to social distancing guidelines and prompted efforts to relax these restrictions [12-14]. Beginning in late April, state governments and the District of Columbia began relaxing the social distancing measures that had, up to that point, successfully slowed the spread of SARS-CoV-2 [3]. Relaxation of such measures is intended to be accompanied by appropriate behavioral practices (e.g., mask wearing and physical distancing) and control measures (e.g., contact tracing and increased availability of testing), so that epidemic control can be maintained [15-21].

However, there has not been a coherent national strategy to promote appropriate behavioral practices, nor has an effective control infrastructure been coordinated at the federal level. As a result, recent decisions and timing of relaxing social distancing measures have been challenged. Critical unanswered questions remain about if and how relaxation of social distancing measures can be carried out while effectively maintaining epidemic control. To address this gap in the literature, we abstracted state-level data on the implementation and relaxation of social distancing measures and undertook a longitudinal pretest-posttest comparison group study to determine the extent to which relaxation of social distancing measures has led to a recrudescence in COVID-19 transmission in the U.S.

## Methods

The unit of analysis was each U.S. state (or the District of Columbia). Our primary explanatory variable of interest was time in days, which we divided into two time periods relative to the first date of relaxation of social distancing measures. To do so, we identified all statewide social distancing measures that were implemented and/or relaxed between March 10-July 15, 2020, triangulating data from state government and third-party sources [3] (see Supplementary Appendix for full details of the search and abstracting process). The pre-relaxation observation period included the date of relaxation and began at the later of the date social distancing measures were first implemented in the state or 56 days prior to the date any of the social distancing measures were first relaxed in the state. The post-relaxation period began the day after any of the social distancing measures were first relaxed and extended through to the earlier of the first date any statewide social distancing measure was re-imposed or the date of data abstraction (July 9, 2020).

We then summarized state-specific patterns of implementation and relaxation of statewide social distancing measures. To determine the extent to which states were able to maintain epidemic control after relaxation of social distancing, we used segmented linear regression: we fitted mixed-effects linear regression models, specifying the time-varying, state-specific effective reproduction number (R*_t_*) as our outcome of interest, and a random effect by jurisdiction to account for within-state differences in behavior, policies, or epidemic reporting. R*_t_* corresponds to the expected number of secondary infections generated by each index case. We selected R*_t_* (as estimated by [22]) as our primary outcome to avoid reliance on crude case detection, which is susceptible to biases resulting from differential testing availability and delays in result reporting, both of which are known to be problematic in the U.S. [23, 24]. In contrast, the methods described by [22] estimate disease transmission patterns based on observed SARS-CoV-2-related deaths [6] and thus partially mitigate bias due to testing and reporting patterns. The R*_t_* value for a given day reflects the secondary cases generated by individuals infected on this day. The primary explanatory variables of interest were time in days, relaxation period, a time-by-relaxation-period product term. We also adjusted for day of the week [25] and state-level population density (estimated from 2018 U.S. population data). We conducted a number of sensitivity analyses to probe the robustness of our findings and to further explore patterns of epidemic transmission after measures were relaxed (Supplementary Appendix for full description of secondary analyses). In brief, these analyses included 1) stratifying analyses by type of measures first relaxed; 2) examining days since relaxation of shelter-in-place orders as the primary predictor of interest; 3) using an alternate method of measuring R*_t_* as derived by Hellewell et al [26], 4) fitting a generalizing estimating equations model in place of a linear mixed effects model, 5) varying the pre-relaxation time period to account for state level patterns, 6) specifying log-change in cases and log-change in deaths as our outcome of interest while accounting for incubation periods and times to death after infection [27-29], and 7) considering epidemic size (i.e. cases, deaths, and zenith R*_t_*) and epidemic indicators (current R*_t_* and test positive rate) as predictors of post-relaxation epidemic growth [30].

## Results

Between March 19 and April 7, 2020, all 51 jurisdictions implemented at least one social distancing measure, and most (45 [88%]) implemented a statewide restriction on internal movement (**Supplemental Table 1**). A median of 47 days after social distancing measures were first implemented (interquartile range [IQR], 41-53), between April 20 and June 1, 2020, all 51 jurisdictions relaxed at least one statewide social distancing measure (**Supplemental Figure 1**). The median number of cumulative cases per state on the date of relaxation was 7,883 (IQR, 3,160-23,650). The median number of cumulative COVID-19-attributable deaths per state on the date of first relaxation was 272 (IQR, 113-1056). There was variation in which social distancing measures were initially relaxed. Easing of work restrictions was the most common element of initial relaxation orders in 40 (78%) jurisdictions, followed by reopening of service industry establishments (32 [63%]), reopening outdoor recreational facilities (22 [43%]), rescission of statewide restrictions on internal movement (16 [31%]), and sanctioning of public events (14 [27%]). Only four states (8%) reopened public schools, and none rescinded mandatory quarantines for interstate travel, as part of their initial relaxation orders.

**Figure 1** displays a scatterplot of the estimated *R_t_* for each state by day, before vs. after the first social distancing measures were relaxed, along with a smoothed line derived from locally weighted scatterplot smoothing. During the eight weeks prior to the first date in each state that social distancing measures were initially relaxed, the estimated *R_t_* declined by an average of 0.012 per day (95% confidence interval [CI], -0.013 to -0.012) (Table 1). This period corresponded with a decline from a modeled mean R*_t_* across all states of 1.44 (95% CI, 1.41-1.48) to 0.75 (95% CI, 0.72-0.78). After the first social distancing measures were relaxed, the estimated *R_t_* reversed course and began increasing by an average of 0.019 per day (95% CI, 0.018-0.020) compared with the pre-relaxation period, such that the mean increase in R*_t_* in the post relaxation period was 0.007 units per day (95%CI 0.006 to 0.007), and reached a mean of 1.16 (95% CI, 1.13-1.18) by 56 days after relaxation. If these trends were to continue, the estimated mean *R_t_* would cross 1.50 by approximately 16 weeks after relaxation.

**Figure 1.**
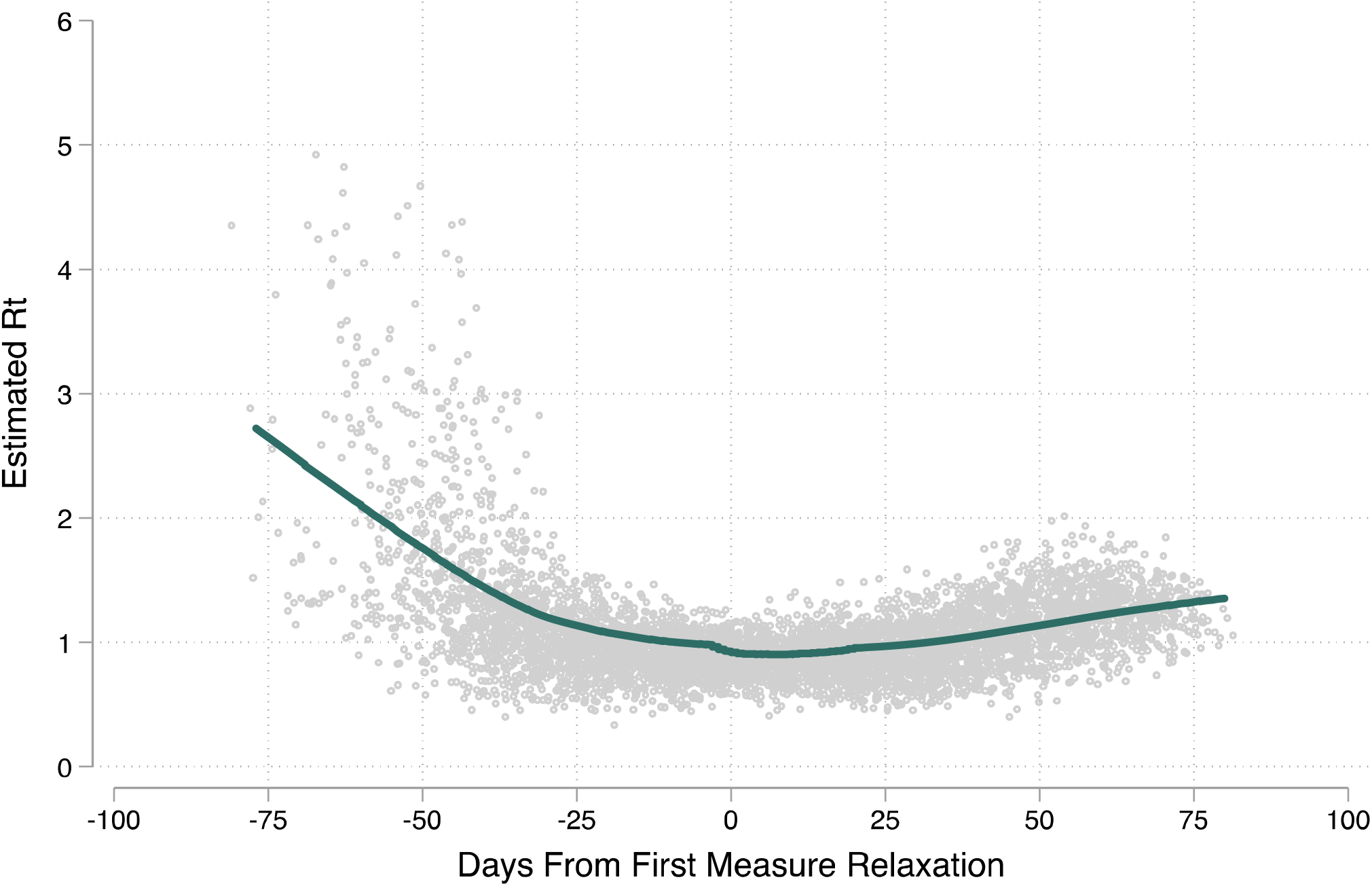
Scatterplot of the estimated *R_t_* for each state by day before and after the first date of relaxation of social distancing measures, along with a smoothed line derived from locally weighted scatterplot smoothing.

**Table 1.** Mixed effects linear regression models for the estimated *R_t_* before vs. after relaxation of social distancing measures

|  | **Coefficient** | **95% Confidence**<br>**Interval** | **P-value** |
| --- | --- | --- | --- |
| Constant term (day prior to relaxation) | 0.761 | 0.728, 0.793 | <0.001 |
| Pre-relaxation period (days relative to relaxation) | -0.012 | -0.013, -0.012 | <0.001 |
| Post-relaxation period intercept | 0.032 | 0.010, 0.054 | 0.005 |
| Time × post-relaxation period (days relative to relaxation) | 0.019 | 0.018, 0.020 | <0.001 |
| Post-relaxation period (days relative to relaxation)<sup>^</sup> | 0.007 | 0.006, 0.007 | <0.001 |

\* Estimates adjusted for day of the week and population density
<sup>^</sup> The post-relaxation term represents the linear combination of the pre-relaxation period and the time × post-relaxation period coefficient

Results were qualitatively similar irrespective of the nature of the first social distancing measures relaxed (Table 2). For each of these regression models, we estimated a statistically significant reversal of R*_t_* from negative to positive after the change from the pre- to post-relaxation period, with estimates ranging from 0.015 (95% CI, 0.013-0.016) for the four jurisdictions that reopened public schools as part of their initial relaxation orders, to a maximum of 0.022 (95% CI, 0.021-0.023) for the 16 jurisdictions that rescinded statewide restrictions on internal movement as part of their initial relaxation orders.

**Table 2.**
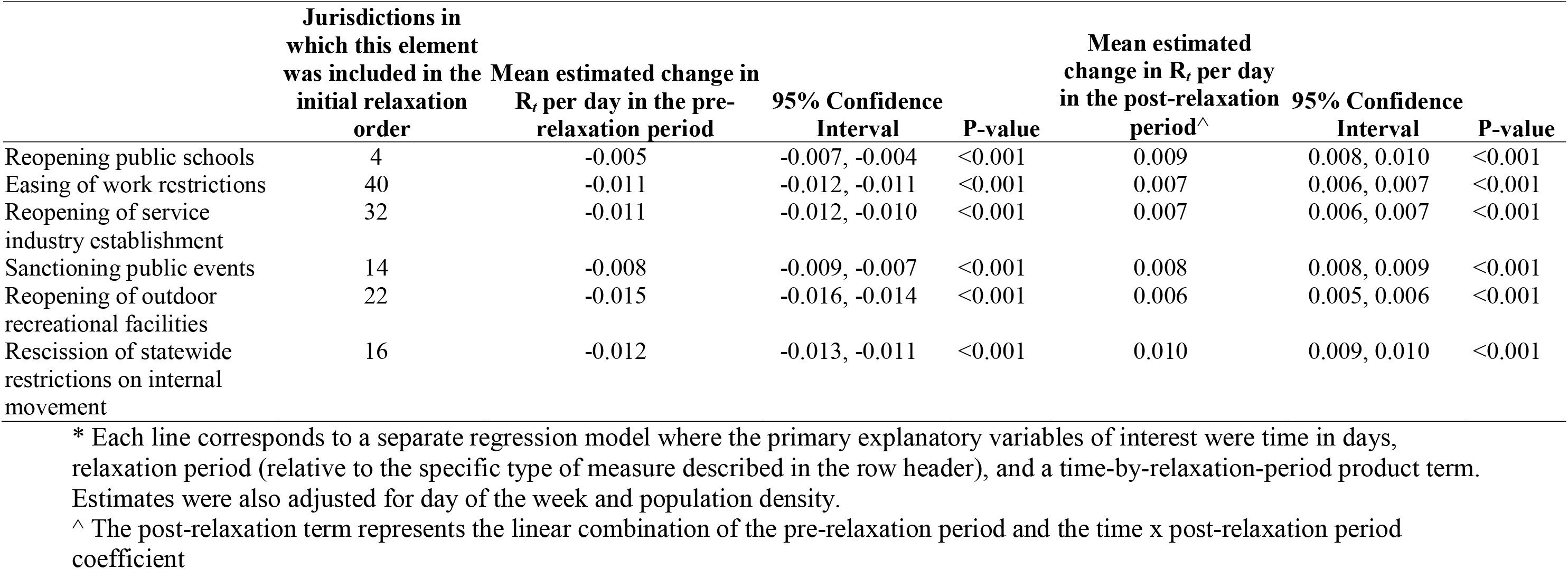
Mixed effects linear regression models for the estimated *R* before vs. after relaxation of social distancing measures, stratified by characteristics of the first relaxation order

Our estimates were robust to several sensitivity analyses (**Supplemental Table 2**). When we redefined the primary explanatory variable of interest as the period before vs. after rescission of statewide restrictions on internal movement, the estimated *R_t_* declined by an average of 0.004 per day (95% CI, -0.005 to -0.004) in the pre-relaxation period. After statewide restrictions on internal movement were lifted, the estimated *R_t_* reversed course and began increasing by an average of 0.013 per day (95% CI, 0.011-0.014) compared with the pre-relaxation period. The estimated regression coefficient on the time-by-post-relaxation period product term was attenuated in magnitude, and slightly attenuated in statistical significance, when we used the *R_t_* estimates based on [26]. The generalized estimating equations specification changed little. Finally, our results remained similar after varying the pre-relaxation period duration from 14 to 42 days, with a downward slope in the pre-relaxation period as we shorted its duration but with persistently significant reversals in the post-relaxation period. When we specified log change in cases and log changes in deaths as the primary outcome of interest, we similarly found a strongly significant conversion from a downward slope to a flattening of the growth rate, corresponding to a consistent rate of increase in cases and deaths.

We inspected state-specific trajectories of *R_t_* during the post-relaxation period to identify correlates of epidemic control after relaxation (**Figure 2**). Forty-four (86%) jurisdictions had established a downward trajectory in *R_t_*, and 46 had achieved an *R_t_* < 1.0 (all jurisdictions excluding Arkansan, Minnesota, Mississippi, Tennessee, and Wisconsin), by the time they had begun relaxing their social distancing measures. However, only 4 states (8%) maintained a negative trend in *R_t_* after relaxation of social distancing: Arkansas, New Jersey, New Mexico, and New York. Nine jurisdictions (18%) maintained an *R_t_* < 1.0 eight weeks following relaxation: Connecticut, the District of Columbia, Maine, Massachusetts, New Hampshire, New Jersey, New Mexico, New York, and Vermont. When we modeled *R_t_* in the post-relaxation period, we found that states with a lower number of cases and lower number of deaths at the time of relaxation, and a higher zenith *R_t_* in the pre-relaxation period, had slightly greater increases in the daily *R_t_* during the post-relaxation period compared with jurisdictions in which the epidemic was more severe prior to or at the time of initial relaxation (**Supplemental Table 3**). Only 9 states (18%) states had achieved a 14-day test-positive rate of <5% at the time of relaxation. Both *R_t_* <0.9 and a test positive rate <5% on the date of relaxation correlated with greater epidemic growth after relaxation (**Supplemental Table 3, Supplemental Figure 2**).

**Figure 2.**
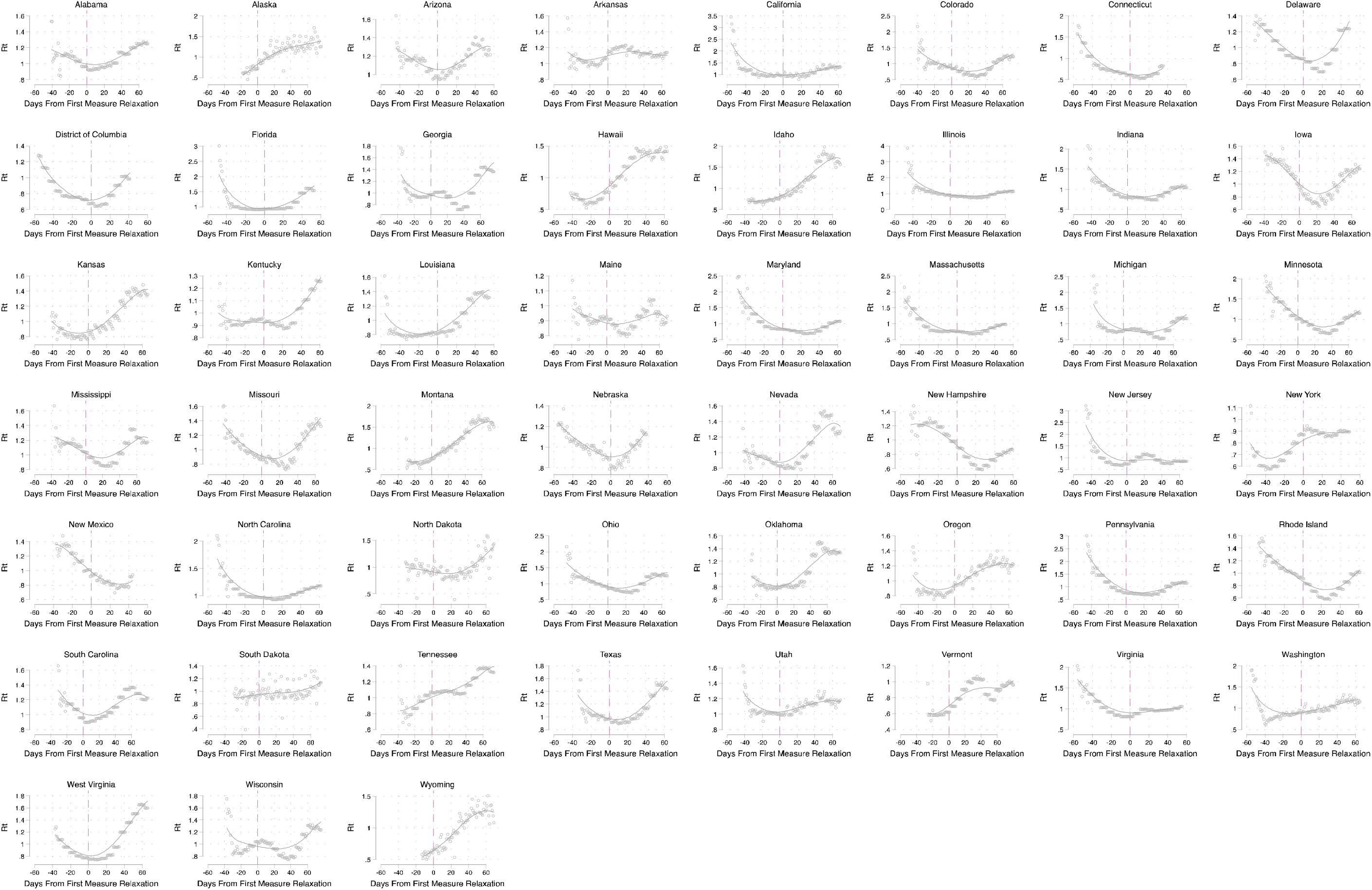
State-specific scatterplots of the estimated *R_t_* by day, before vs. after the first date that social distancing measures were relaxed, along with a smoothed line derived from locally weighted scatterplot smoothing.

## Discussion

In this national study observing the COVID-19 epidemic during the period April - July 2020 in the U.S., we found that relaxation of statewide social distancing measures was associated with a reversal of the downward trend in transmission of SARS-CoV-2 that had been achieved after these measures were implemented. In all but nine states, the reversal returned the estimated *R_t_* back above 1.0 within eight weeks of the initial relaxation of social distancing measures. These patterns were apparent irrespective of the specific kinds of social distancing measures that were relaxed and irrespective of key indicators of epidemic severity (e.g., test positivity rate) that have been heretofore used by many jurisdictions as one indicator to guide relaxation decisions [30]. Our findings, in combination with prior data noting the strong and significant effect on epidemic interruption after implementation of measures [2-7], should motivate policymakers to reconsider the rapid pace at which states are reopening their economies. Furthermore, in the states that are currently experiencing a recrudescence of SARS-CoV-2 transmission, strong consideration should be given to the re-imposition of social distancing measures so that new infections of COVID-19 do not overwhelm the local health care system. Intermittent social distancing regulations may be necessary to control the COVID-19 epidemic in the U.S. until more effective treatments or an effective vaccine become available and achieve widespread dissemination in the population [21].

Little data exist to inform the U.S. exit strategy from its current state of social distancing measures. The city of Wuhan in Hubei Province, China was the first to enter a regime of strict social distancing (beginning January 23, 2020), but the relaxation of these measures has not resulted in a resurgence of SARS-CoV-2 transmission [31]. Within two months, the Chinese government was able to achieve the milestone of having five consecutive days in which there were no new locally transmitted cases in the country [32]. Modeling studies, however, suggest that stringent social distancing should be maintained for longer than the median duration observed in the U.S. and that social distancing should be treated as a strategy for suppressing SARS-CoV-2 transmission so that additional non-pharmaceutical interventions (e.g., contact tracing and increased availability of testing) can be deployed [15-20, 33, 34]. Moreover, deconfinement should occur gradually and, in the setting of well-connected jurisdictions (e.g., countries in Europe or states in the U.S.), should be coordinated across jurisdictions to maximize the probability of successful deconfinement [35].

We found that states with more severe epidemics at the time social distancing measures were relaxed had reduced post-relaxation epidemic growth, compared with states that had experienced smaller epidemics. These differences were small in magnitude but precisely estimated. This finding suggests that individuals living in states with large epidemics might be more likely to maintain social distancing even after local orders are formally lifted. These results are in keeping with data from survey respondents in New York City and Los Angeles during the peak of their epidemics, who showed near unanimous support for such measures in locations which had suffered severe epidemics [36].

We also explored whether epidemic indicators can be used to guide re-opening. One such indicator, recommended by the World Health Organization and multiple U.S. states, is a test positive rate of less than 5% over 14 days [30]. Although a handful of states met this recommended threshold when social distancing measures were relaxed, those that did unexpectedly experienced a greater increase in epidemic transmission following relaxation. Similarly, states with a lower *R_t_* at the time of relaxation saw a faster subsequent increase. In summary, in the U.S. there appeared to be a paradoxical inverse correlation between indicators of epidemic control at the time social distancing measures were relaxed and subsequent trajectories of SARS-CoV-2 transmission. Indeed, these data might suggest that if the public over-interprets government-issued relaxation of measures as indicators of epidemic control, and as a result disregards non-mandated social distancing practices (i.e. mask wearing and physical distancing), a counter-intuitive worsening might follow their rescission.

The primary limitation of our analysis is the potential for confounding by phenomena that occur simultaneously with relaxation of social distancing measures and also influence the trajectories of *R_t_*. For example, if a social movement supportive of a reopening agenda [37] advocates for relaxation of state measures and independently influences social distancing behaviors [12, 14], we could misclassify the effect seen to be due to the measure relaxation (in place of the social movement). However, we found similar effect sizes and time-specific reversal in *R_t_* trends across most states, and irrespective of political and demographic characteristics. Moreover, because relaxation took place on across a wide range of dates, confounders who would have to occur independently across multiple states and similarly vary with relaxation of state measures.

Notwithstanding this potential limitation, our findings suggest that suppression of SARS-CoV-2 after removal of statewide social distancing regulations has failed. Robust surveillance programs are needed so that, should the observed trends continue, state and local public health policy makers can continuously evaluate the stringency of social distancing measures required to prevent subsequent epidemic surges [15-20, 33, 34]. Considering the current projected timelines for vaccine development [38], and low levels of cumulative population infection even in countries that have experienced severe epidemics [39, 40], thoughtful public health leadership will be needed to ensure that COVID-19-attributed mortality is maximally prevented.

## Data Availability

The data on social distancing policies are contained within the Supplementary Appendix Table. The data on COVID-19 cases and deaths are publicly available from https://github.com/nytimes/covid-19-data.

